# The Sidelines Effect on Minds: The Impact of Injury on Mental Health in National Collegiate Athletic Association Athletes

**DOI:** 10.1101/2023.08.03.23293610

**Authors:** Ruth A. Axton, Nancy A. Crowell, Daniel J. Merenstein

**Affiliations:** Department of Human Science, Georgetown University; School of Nursing, Georgetown University

## Abstract

Injury and mental health are prevalent topics in among NCAA Division 1 (DI) student-athletes; however, there is limited literature regarding the impact of injury on student-athletes’ mental health. This study examined the impact of injury on social support and self-stigma and the relationship between athletic identity and self-stigma. A convenience sample survey yielded 101 responses from DI student-athletes. The survey was composed of demographic questions, Sarason Social Support Questionnaire, Self-Stigma for Seeking Self-Help Scale, and Athletic Identity Measurement Scale. There was no significant difference between injured and non-injured athletes in the mean scores of social support (t test p= 0.69) or self-stigma (t test p =0.92). There was a nonsignificant weak correlation between athletic identity and self-stigma. The results reveal that self-stigma is prevalent within this community, with an average score of 23.34 for all respondents. These conclusions can aid mental health professionals in providing support and education to student-athletes.

---

Recent studies have shown that one in four NCAA DI student-athletes report depressive symptoms, as compared to 13.1%. prevalence in traditional college students (Wolanin et al., 2016 & NIMH, 2019). Increased prevalence of mental health concerns, highlighted by the loss of five student athletes by suicide in 2022 (Mazzella, 2023), and the concern for student athlete mental health following the COVID-19 pandemic (*NCAA Student-Athlete Well-Being Study*) has pushed the topic to the forefront of conversation. Among other factors, mental health in athletes is moderated by social support, self-stigma for seeking help, and Athletic Identity. (Antoniak et al., 2022; Hefner & Eisenberg, 2009; Hilliard et al., 2022; Nippert & Smith, 2008; Rice et al., 2022; Sullivan et al., 2022). Injury is a common consequence of athletic performance and one that isolates student-athletes from their team, which could challenge their social support network or athletic identity.

## Social Support

Social support is a person’s network of people or groups that one can depend on for support (Sarason *et. al*, 1983). Student-athletes draw social support from teammates, coaches, athletic training staff, and strength and conditioning staff (Storch et al, 2005). Social support has been shown to be a protective factor against student-athlete burnout and enhanced gratitude and sports satisfaction (Gabana et al, 2017), whereas lack of support has been shown to be a barrier to using mental health services (Gulliver *et. al,* 2012). In injured student-athletes, the source of social support for student-athletes tends to shift more to athletic training staff as they spend time away from their team (Barefield, 1997).

## Self-Stigma

Self-stigma can be defined as internalizing the public attitudes surrounding mental health and suffering numerous negative consequences as a result (Corrigan, 2013). High levels of self-stigma are a barrier to the use of mental health services (Gulliver et al, 2012).

## Athletic Identity

Athletic Identity is defined as the degree and strength that one identifies with being an athlete. Athletic identity has been shown to be a predictor of an athlete’s mental toughness (Altıntaşa and Keleceka, 2017). Mental toughness is a concept in sports that athletes should be able ignore all outside factors to physically perform (Gucciardi et al, 2017). It is possible that advocating for mental toughness could lead to student-athletes lacking support in sharing their mental health concerns, which is correlated with decreased use of psychological service and increased self-stigma (Nam *et. al,* 2013). Other sources highlight that mental toughness may be positively correlated with mental health (Gucciardi et al, 2017). Athletic identity has also been shown to have a negative correlation with attitudes about seeking mental health support (Weatherhead, 2015 & Bauman, 2016).

The objective of this study was to examine the effect of injury on the social support and self-stigma of NCAA DI student-athletes and determine the relationship between self-stigma and athletic identity. We hypothesize that injured student-athletes will have lower levels of social support than non-injured student-athletes, self-stigma will be higher in non-injured athletes, and student-athletes with higher athletic identity will show higher levels of self-stigma.

## Method

### Participants

Participants were NCAA DI student-athletes over the age of 18 from 6 NCAA DI universities. The sample yielded 126 initial responses.

### Material

The IRB Approved survey (ID:00004625) collected non-identifying demographic questions (age, gender identity, sport played, year in school, injury status, and previous mental health diagnoses) and three validated instruments (Sarason et al, 1983 & Cohen et al, 1985).

Social support will be measured using the Sarason Social Support Questionnaire, specifically, the Interpersonal Support Evaluation List (shortened version) as its use has been validated in university students and the shortened version allows the survey to be accessible to larger groups of athletes. (Cohen et al, 1985). The Sarason Social Support Questionnaire is a 12- item instrument which evaluates the number of social supports and the satisfaction with social support that is available. Participants rate on a 1-4 likert scale ranging from “Definitely False” to “Definitely True.”

Self-stigma in student-athletes will be measured using the Self-Stigma of Seeking Self-Help Scale (SSOSH), which has been shown to have good reliability, unidimensional factor solution, and has been validated in revealing a positive correlation with a tendency to conceal personal information (Vogel et al., 2006).Self-stigma for Seeking Self-Help Scale is a 10-item instrument that evaluates the participants’ perception that seeking help would harm their self-confidence or worth as a person (Vogel et al, 2006). Participants rate instrument items on a scale of 1-5 likert scale ranging from “Strongly Disagree” to “Strongly Agree.”

The Athlete Identity Measurement Scale is a 10-item instrument used to identify how much participants identify with the athlete role (Brewer, 1993). The scale uses a 1-7 likert scale with answers ranging from “Strongly Disagree” to “Strongly Agree.” The survey was administered through Qualtrics, and responses were collected during a 4-week period from January 21st, 2022 to February 21st, 2022. A copy of the survey is included in Appendix A.

### Procedures

The study was sent to a convenience sample of student-athletes on various teams at six NCAA DI institutions. Consent for the study was imbedded within the survey. Responses were collected during January and February of 2022.

### Statistical Analysis

Survey data on social support, self-stigma, and athletic identity were analyzed using STATA. The difference in mean scores for social support and self-stigma for injured and non-injured student-athletes was analyzed using a t test approach. The relationship between athletic identity and Self-Stigma was analyzed using a correlation coefficient.

## Results

A total of 126 responses were collected. Responses without complete answers to the Sarason Social Support Questionnaire, Self-Stigma for Seeking Self-Help Scale, or Athletic Identity Measurement Scale were removed and 101 responses were analyzed.

***Table 1*** describes the demographics of the survey respondents.

**Table 1.** Demographics.

|  | **Injury** | **Non-Injury** |
| --- | --- | --- |
| **Total** | 64 | 37 |
|  | **M (SD)** | **M(SD)** |
| **Age** | 20.43(1.28) | 19.59 (1.39) |
| **Class** | **Total** | **Total** |
| Freshman | 10 | 14 |
| Sophomore | 17 | 12 |
| Junior | 11 | 5 |
| Senior | 24 | 6 |
| Graduate | 2 | 0 |

| **Gender Identity** |  |  |
| --- | --- | --- |
| Female | 47 | 29 |
| Male | 17 | 8 |
| **Sports Team** |  |  |
| Field Hockey | 6 | 5 |
| Baseball | 7 | 3 |
| Women's Rowing | 7 | 12 |
| Women's Lacrosse | 7 | 3 |
| Swimming | 3 | 2 |
| Men's Soccer | 1 | 0 |
| Women's Soccer | 0 | 1 |
| Women's Tennis | 1 | 1 |
| Men's T&F/XC | 1 | 2 |
| Volleyball | 1 | 1 |
| Men's Lacrosse | 0 | 1 |

### Social Support and Injury

There was no significant difference between the responses to the Sarason Social Support Questionnaire for those who were injured and those who were not injured (t(93) = 0.33 p = 0.69). (Figure 1)

**Figure 1.**
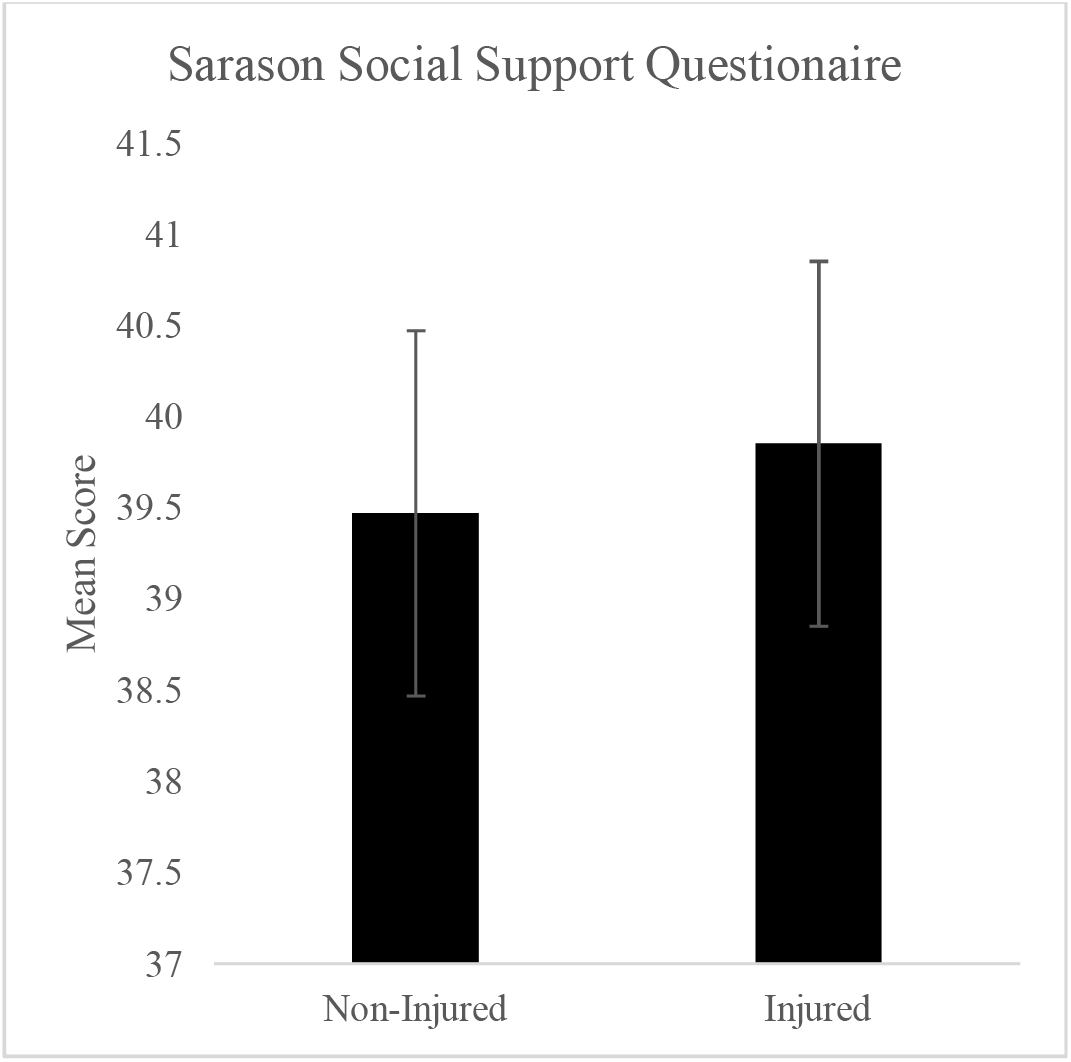
Responses from Sarason Social Support Questionnaire *Note*: Mean Scores for Injured (39.85246) and non-injured (39.47059) student-athletes on the Sarason Social Support Questionnaire.

### Self-Stigma and Injury

There was no significant difference between the responses to the Self-stigma for Seeking Self-Help for those who were injured and those not injured (t(99) = 1.40, p = 0.92). (Figure 2)

**Figure 2.**
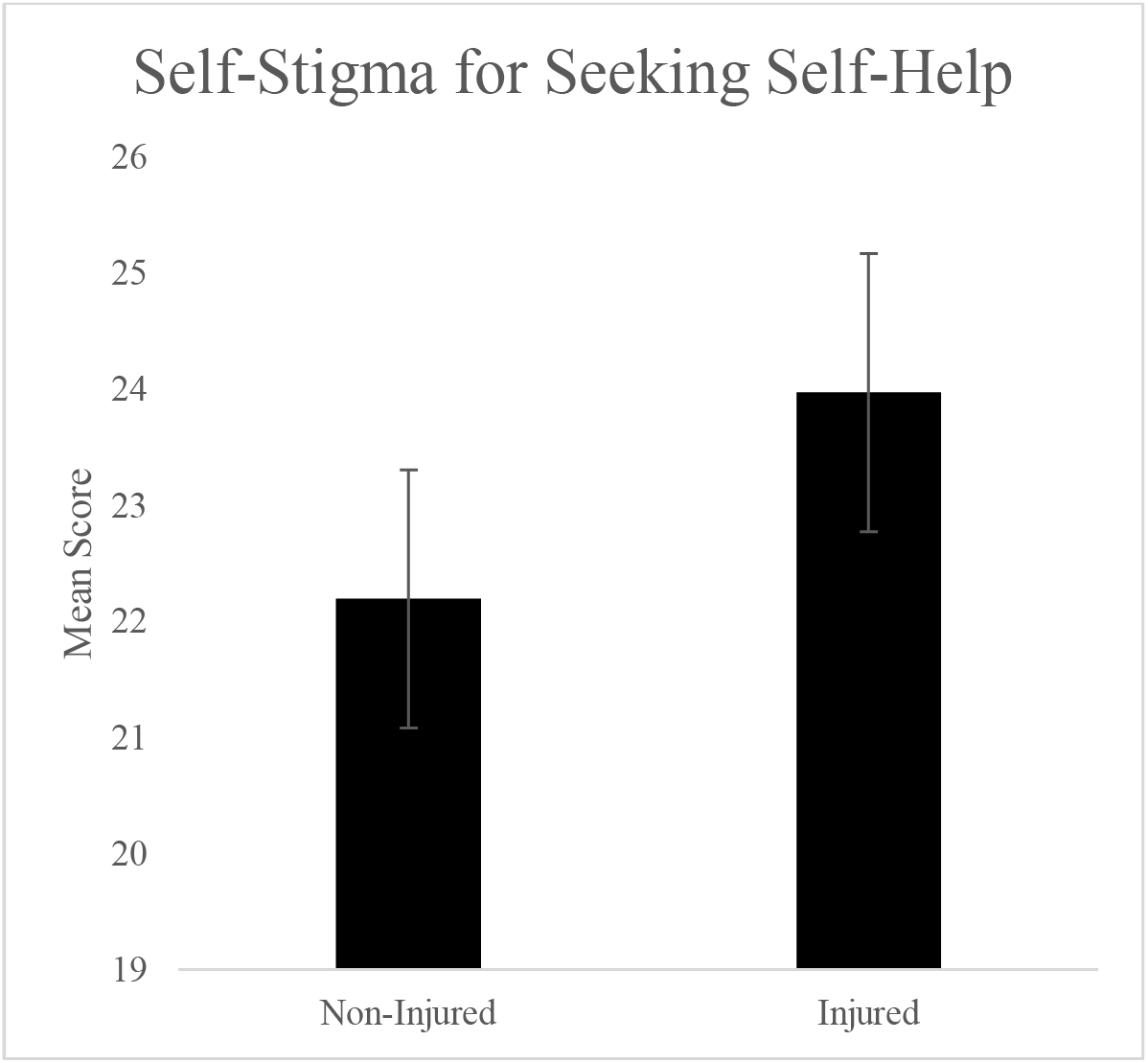
Responses from the Self-Stigma for Seeking Self-Help Questionnaire *Note*: Mean scores for Injured (23.97) and non-injured (22.19) student-athletes on the Self-Stigma for Seeking Self-Help scale.

### Self-Stigma and Athletic Identity

There was a weak relationship (r=0.20) between athletic identity and self-stigma for seeking self-help, which was not statistically significant (p = 0.051).

**Figure 3.**
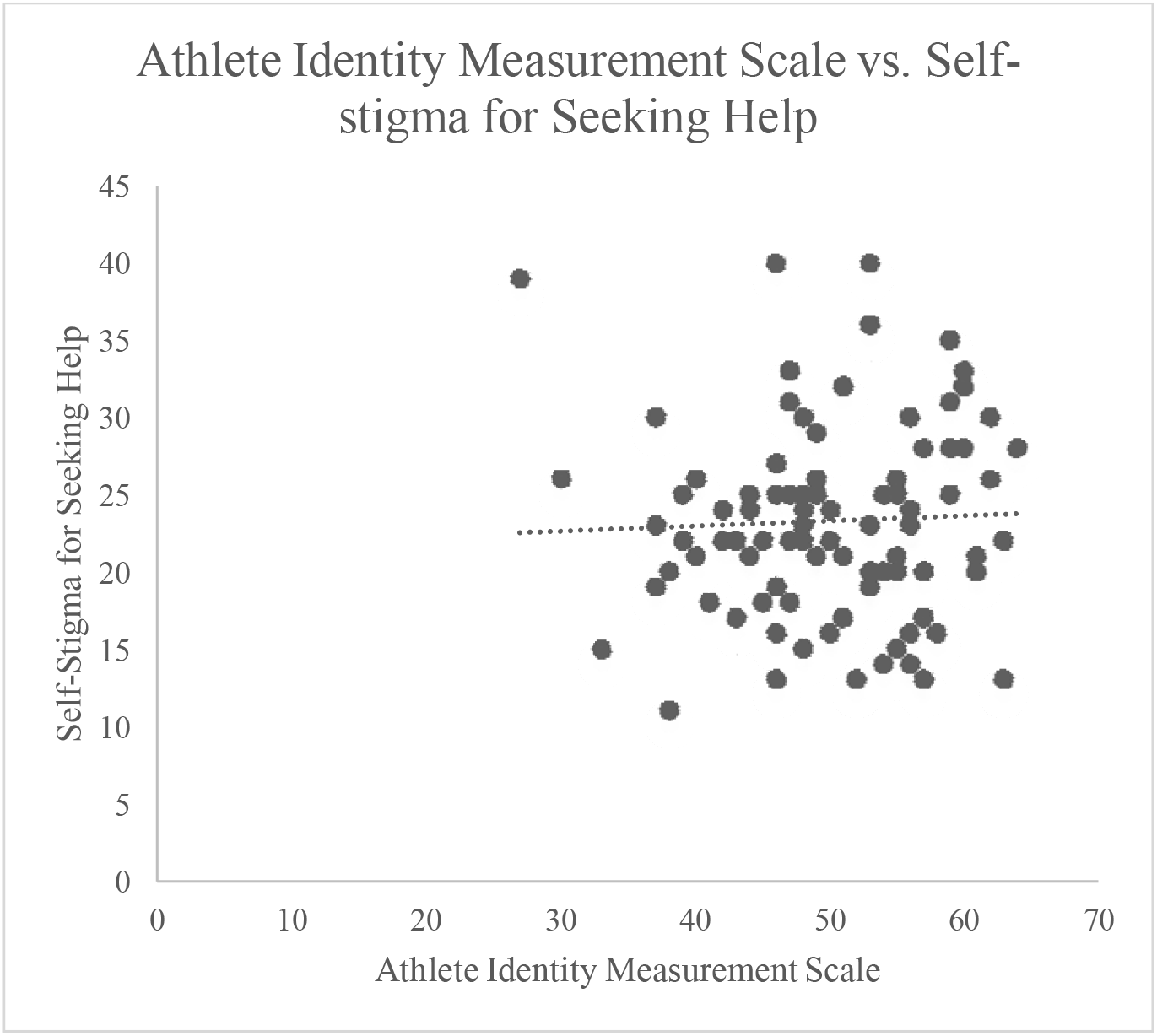
Correlation between Athlete Identity and Self-Stigma *Note:* Correlation between scores from Athletic Identity Measurement Scale and Self-Stigma for Seeking Self-help, trendline (y = 15.2 + 0.16x) and correlation coefficient (R = 0.20).

## Discussion

### Social Support

It was hypothesized that injured student-athletes would have lower levels of social support compared to their non-injured counterparts. However, results revealed no significant difference. The lack of differentiation between these two groups may be a result of the change in the support system for injured student-athletes from teammates to other support staff (athletic trainers, doctors, etc.). Additionally, the structure of collegiate athletics may lessen the effect of injury-related isolation as 53 and 49 percent of male and female DI student-athletes, respectively, are roommates with other student-athletes (*Come and Knock on Our Door)*. This could provide an additional amount of support which is not disrupted by injury status.

## Self-Stigma

It was hypothesized that injured student-athletes would have higher levels of self-stigma than non-injured athletes. No significant difference was found within our population. While there was no data suggesting a relationship between injury and self-stigma, self-stigma was notable in both groups. The presence of self-stigma within student-athletes can be detrimental to the overall mental health of student-athletes, leading to discomfort seeking mental health services (Bird et. al, 2018 & Hillard et. al, 2019). The presence of self-stigma reflected on the SSSH scale within both the injured and non-injured groups, 23.97 and 22.19 respectively, highlights that it is necessary to address this stigma in all athletes. Over time high levels of public stigma surrounding mental health care are correlated with a higher level of self-stigma (Vogel et al, 2013). This correlation is indicative of an opportunity to lower self-stigma in future student athletes by focusing on public acceptance of mental health treatment. This acceptance could be facilitated by increased education on mental health throughout all levels of athletic departments.

The presence of self-stigma within athletes could also be a manifestation of label avoidance. This is the facet of stigmatization where people avoid seeking mental health care to avoid receiving a stigmatizing label (Corrigan & Wassel, 2008). It is possible that athletes could avoid seeking mental health care to not affect their ability to participate or their perceived ability to perform in their sport. The prevalence of label avoidance in the athletic population would need to be individually measured but could also benefit from the increase in education and acceptance from coaches, staff, and administrators within athletic departments.

### Limitations

A major limitation of this study was the potential influence of the Coronavirus pandemic on survey responses. Student-athletes and college students were more isolated from their peers during the pandemic due to the cancellation of sports and in-person classes, (Giovenco et al., 2022 & *NCAA Student-Athlete Well-Being Study*) which may have impacted responses to the SSSQ. Additionally, the separation of student-athletes from their sports and teammates during this time could have impacted responses to the AIMS. The time of year when the survey was distributed is in correlation with a heightened prevalence of Seasonal Affective Disorder (SAD). This may have influenced all results to be lower than those when SAD prevalence is lower. To control for this variable, student-athletes could be surveyed at different points throughout the year.

Another limitation was the small sample size which may prevent the generalizability of the results to all NCAA DI student-athletes. Because a large percentage of respondents were from women, further research with a larger population is required to generalize these results to all of DI student-athletes.

### Future Research

Future projects would benefit from looking at social support and self-stigma in student-athletes compared to traditional college students. This would help to elucidate if the team environment is helpful in creating a network of social support for student-athletes that is not present for traditional college students. Research could also focus on the relationship between mental toughness and athletic identity and how that impacts student-athlete self-stigma. Mental toughness was found to be a factor correlating with positive mental health in athletes, and it would be important to identify if that also correlates with lower levels of self-stigma.

Another future direction could include reconducting this survey after the pandemic has ended. This could reveal a comparison between results from during and after the pandemic showing the effects it had on student-athletes. It could also elucidate the effects of the increased conversation surrounding student-athlete mental health in the recent years.

## Conclusion

This study was successful in investigating the effect of injury on social support, self-stigma, and athletic identity. Although no statistically significant difference was found between injured and non-injured athletes for either variable and no significant correlation was found between athletic identity and self-stigma, these results provide important insight for athletic counseling. There was a presence of self-stigma in this group of student-athletes highlighting the need for continued mental health education and support for both injured and non-injured student-athletes. Additionally, the weak correlation between athletic identity and self-stigma highlights that self-stigma is not only found in athletes with high athletic identity and efforts to reduce stigma can be applied to all athletes.

## Data Availability

All data produced in the present study are available upon reasonable request to the authors

## Appendix A

Included below is the IRB Approved Survey given to participants.

Demographics:

1. Age:
2. Gender identity
3. Year in School
4. Sport played
5. Have you been injured while competing in college athletics?
6. If yes, what was the nature of your injury?
7. Did the injury preclude you from competing in sport for at least one day?
8. Have you ever been diagnosed with the following:

1. Anxiety
2. Depression
3. OCD
4. Personality Disorder
5. Eating Disorder
6. Other Mental Health Diagnoses
7. None

Please use the 5-point scale to rate the degree to which each item describes how you might react in this situation.

1= Strongly Disagree

2 = Disagree

3 = Agree and Disagree Equally

4 = Agree

5 = Strongly Agree

1. I would feel inadequate if I went to a therapist for psychological help.
2. My self-confidence would NOT be threatened if I sought professional help.
3. Seeking psychological help would make me feel less intelligent.
4. My self-esteem would increase if I talked to a therapist.
5. My view of myself would not change just because I made the choice to see a therapist.
6. It would make me feel inferior to ask a therapist for help.
7. I would feel okay about myself if I made the choice to seek professional help.
8. If I went to a therapist, I would be less satisfied with myself.
9. My self-confidence would remain the same if I sought professional help for a problem I could not solve.
10. I would feel worse about myself if I could not solve my own problems.

For each statement circle "definitely true" if you are sure it is true about you and "probably true" if you think it is true but are not absolutely certain. Similarly, you should circle "definitely false" if you are sure the statement is false and "probably false" if you think it is false but are not absolutely certain.

1 = definitely false

2 = probably false

3 = probably true

4 = definitely true

1. If I wanted to go on a trip for a day (for example, to the country or mountains), Iwould have a hard time finding someone to go with me.
2. I feel that there is no one I can share my most private worries and fears with.
3. If I were sick, I could easily find someone to help me with my daily chores.
4. There is someone I can turn to for advice about handling problems with my family.
5. If I decide one afternoon that I would like to go to a movie that evening, I could easily find someone to go with me.
6. When I need suggestions on how to deal with a personal problem, I know someone I can turn to.
7. I don’t often get invited to do things with others.
8. If I had to go out of town for a few weeks, it would be difficult to find someone who would look after my house or apartment (the plants, pets, garden, etc.).
9. If I wanted to have lunch with someone, I could easily find someone to join me.
10. If I was stranded 10 miles from home, there is someone I could call who could come and get me.
11. If a family crisis arose, it would be difficult to find someone who could give me good advice about how to handle it.
12. If I needed some help in moving to a new house or apartment, I would have a hard time finding someone to help me.

Instructions: Please use the 7-point scale to rate the degree to which each item describes how you feel about the following statements.

1= Strongly Agree

2 = Agree

3 = Agree Somewhat

4 = Neither Agree or Disagree

5 = Disagree Somewhat 6= Disagree

7 = Disagree Strongly

1. I consider myself an athlete.
2. I have many goals related to sport.
3. Most of my friends are athletes.
4. Sport is the most important part of my life.
5. I spend more time thinking about sport than anything else.
6. I need to participate in sport to feel good about myself.
7. Other people see me mainly as an athlete.
8. I feel bad about myself when I do poorly in sport.
9. Sport is the only important thing in my life.
10. I would be very depressed if I were injured and could not compete in sport.

